# Making an equal system equitable: Proposing a sex-adjusted MELDNa score for liver transplantation allocation

**DOI:** 10.1101/2021.07.12.21260196

**Authors:** Julia M. Sealock, Ioannis A. Ziogas, Zhiguo Zhao, Fei Ye, Sophoclis P. Alexopoulos, Lea Matsuoka, Guanhua Chen, Lea K. Davis

**Affiliations:** Division of Genetic Medicine, Department of Medicine, Vanderbilt University Medical Center, Nashville, TN, 37232, USA; Vanderbilt Genetics Institute Vanderbilt University Medical Center Nashville, TN, 37232, USA; Division of Hepatobiliary Surgery and Liver Transplantation, Vanderbilt University Medical Center, Nashville, TN, 37232, USA; Department of Biostatistics, Vanderbilt University Medical Center, Nashville, TN, 37232, USA; Department of Biostatistics and Medical Informatics, University of Wisconsin-Madison, Madison, WI, 53706, USA; Department of Molecular Physiology and Biophysics, Vanderbilt University Medical Center, Nashville, TN, 37232, USA; Department of Psychiatry and Behavioral Sciences, Vanderbilt University Medical Center, Nashville, TN, 37232, USA; Department of Medicine and Biomedical Informatics, Vanderbilt University Medical Center, Nashville, TN, 37232, USA

**Keywords:** sex characteristics, liver transplant, electronic health records, MELDNa scores, biomarkers

## Abstract

**Background & Aims:** Liver allocation is determined by the model for end-stage liver disease (MELD), a scoring system based on four laboratory measurements. During the MELD era, sex disparities in liver transplant have increased and there are no modifications to MELD based on sex. We use data from electronic health records (EHRs) to describe sex differences in MELD labs and propose a sex adjustment.

**Methods:** We extracted lab values for creatinine, International Normalized Ratio of prothrombin rate, bilirubin, and sodium from EHRs at Vanderbilt University Medical Center (VUMC) and the All of Us Research Project to determine sex differences in lab traits. We calculated MELDNa scores within liver transplant recipients, non-transplanted liver disease cases, and non-liver disease controls separately. To account for sex differences in lab traits in MELDNa scoring, we created a sex-adjusted MELDNa map which outputs adjusted female scores mapped to male scores of equal liver disease severity. Using waitlist data from the Liver Simulated Allocation Modeling, we conducted simulations to determine if the sex-adjusted scores reduced sex disparities.

**Results:** All component MELDNa lab values and calculated MELDNa scores yielded significant sex differences within VUMC (n=623,931) and All of Us (n=56,715) resulting in MELDNa scoring that disadvantaged females who, despite greater decompensation traits, had lower MELDNa scores. In simulations, the sex-adjusted MELDNa score modestly increased female transplantation rate and decreased overall death.

**Conclusions:** Our results demonstrate pervasive sex differences in all labs used in MELDNa scoring and highlight the need and utility of a sex-adjustment to the MELDNa protocol.

**Lay Summary:** Liver transplant waitlist position is determined by a score called MELDNa, which is calculated using four laboratory values. Once on the waitlist, males are more likely to receive a transplant, while females are more likely to die or be removed due to illness. We demonstrate that all four laboratory values in the MELDNa score show significant sex differences that disadvantage females in liver transplant. We created a sex-adjusted score that increases female transplantation rate and decreases death among both sexes in simulations.

## Introduction

In 2002, the Organ Procurement and Transplantation Network (OPTN) adopted the model for end-stage liver disease (MELD) scoring system to allocate livers based on objective medical criteria. MELD scores were initially calculated from three laboratory values, creatinine, international normalized ratio of prothrombin rate (INR), and bilirubin^1^. The MELD allocation system is based on the “sickest first” principle whereby individuals with the highest scores have priority access to organs. In some cases, MELD does not adequately capture the severity of illness, such as hepatocellular carcinoma, hepatopulmonary syndrome, and portopulmonary hypertension^2^. Individuals with these diagnoses receive exception points to increase their listing MELD scores and decrease their time to transplant. Additionally, other factors can affect MELD’s mortality prediction^2^, including hyponatremia, nutritional status, and sex^3,4^. These effects on MELD’s prediction led to several proposed modifications to MELD scoring^2^, including the sodium-adjusted MELD score^5,6^ (MELDNa) which has replaced the original MELD in clinical practice. Notably, sex disparities in liver transplant have widened in the MELD era^7,8^, however, no modifications to MELD based on sex have been accepted.

Sex differences in liver transplant are well documented. Females spend longer on the waitlist^8,9^, are more likely to die or become too sick for transplantation while on the waitlist^3,8,10–12^, and are less likely to receive MELD exception points than males^13^. Previous studies attempted to explain the sex disparity in liver transplants by investigating factors such as size mismatches between donor-recipient pairs, geographic disparities, and lower creatinine levels in females. After controlling for estimated liver size^12,14^ or height^15^, sex disparities in transplant were reduced, but not eliminated. Likewise, controlling for geographic disparities did not ameliorate the sex disparity in receiving a liver transplant^12^. Several studies postulated that decreased body size in females leads to lower creatinine levels which in turn disadvantages females in MELD scoring^3,16,17^. Indeed, males tend to be listed with higher creatinine, estimated glomerular filtration rate (eGFR), and MELD scores than females^3,4,16–18^, but replacement of creatinine with eGFR did not improve the MELD model in females^3,11,18^. The lack of explanation from previous studies suggests no single factor can fully explain the sex difference in liver transplant, but rather a constellation of differences exist between males and females, leading to the observed disparity.

The majority of previous studies investigating sex differences in liver transplant utilized data from the United Network for Organ Sharing (UNOS) database, which allows for analysis of demographics, cause of liver disease, MELD scores at listing, comorbidities, and outcomes. However, the UNOS database does not allow for investigation of sex differences at the population level. Electronic health records (EHRs) store longitudinal information on the health and clinical care of individuals, including diagnoses, procedures, medications, and laboratory test results. Rather than conducting studies in traditionally collected cohorts, EHRs enable research on the entire population of a healthcare system, which can increase sample size, reduce bias based on ascertainment, and increase generalizability. In this study, we leverage EHR data from a single tertiary care medical center and a multisite initiative, the All of Us Research Program^19^, to investigate sex differences in the (1) lab traits composing MELDNa scores, (2) calculated MELDNa scores, and (3) number of liver decompensation traits. To form a complete picture of the scope of sex differences affecting MELD we included individualsat a range of clinical endpoints from healthy controls to liver disease to transplant cases. Finally, we derive a sex-adjusted MELDNa score and test its ability to reduce sex disparities in simulated liver transplant waitlist data to ensure applicability to a transplant-specific sample.

## Methods

### Data Source

Vanderbilt University Medical Center (VUMC) is a tertiary care center that provides inpatient and outpatient care in Nashville, TN. The VUMC EHR was established in 1990 and includes data on billing codes from the International Classification of Diseases, 9^th^ and 10^th^ editions (ICD-9 and ICD-10), Current Procedural Terminology (CPT) codes, laboratory values, reports, and clinical documentation. The de-identified mirror of the EHR, numbers more than 3.2 million patient records. This study was deemed non-human subjects research by the VUMC Institutional Review Board (IRB#172020).

### Study Sample

To maximize the density of records across our study sample, we implemented a data floor heuristic (at least 5 ICD codes received over a period of at least 3 years) to select individuals who regularly utilize VUMC for their primary care. Because liver cancer and renal dialysis affect MELDNa scores, individuals with the presence of the liver cancer ICD codes (155-155.2, C22-C22.9) or dialysis CPT codes (90935-90999) were excluded, resulting in 623,931 individuals available for subsequent analyses (Supplementary Table 1). Liver transplant cases were defined by the presence or absence of the liver transplant CPT code, 47135 (N=601). The remaining sample was further stratified by liver disease status, determined by the presence of at least two component chronic liver disease ICD codes (Supplementary Table 2), indicative of individuals who might qualify for a transplant but have not yet received one (N=24,921). Individuals with only one liver disease code were excluded to reduce potential false positive cases. The remaining sample was classified as “healthy controls” (N=598,409). In this study, “healthy control” status refers to the absence of liver disease. EHRs are not linked to the liver transplant waitlist system, therefore we were not able to use waitlist-specific information, such as listing date, dual listing status, or de-listing status.

## Statistical Analysis

### Lab Values

MELDNa component labs (creatinine, INR, bilirubin, and sodium) were extracted from the EHR and filtered for observations more than eight standard deviations from the group sample mean, indicative of data entry error or biologically implausible values. Given the longitudinal nature of the EHR, we selected the median and maximum of each lab value per individual. In liver transplant cases, we selected the median and maximum lab values prior to transplant, as defined as the date of liver transplant CPT code (47135). In liver disease cases and in controls, the median and maximum lab values across the entire medical record were selected (Supplementary Table 3). We applied t-tests to determine the differences in lab values between males and females within the entire sample, within healthy controls, within the liver disease sample, and within the liver transplant sample.

### OPTN MELDNa Scores

OPTN MELDNa scores were calculated using all available eight SD filtered creatinine, INR, bilirubin, and sodium values. To most closely estimate the clinically relevant scores, we required that all four lab values contributing to MELDNa score calculations were recorded on the same date. For liver disease cases and healthy controls, median and maximum OPTN MELDNa (MELDNa_median_ and MELDNa_max_) across the entire medical record were used for subsequent analyses. For liver transplant cases, OPTN MELDNa_median_ and MELDNa_max_ prior to liver transplant CPT code were used in order to more closely estimate pre-transplant liver disease severity. Differences in OPTN MELDNa scores were assessed between males and females across the entire sample and within liver transplant, liver disease, and control definitions, using t-tests.

### Decompensation Measurements

Decompensation traits were defined using ICD 9 and 10 codes for hematemesis, gastrointestinal hemorrhage, ascites, jaundice, and hepatic encephalopathy (Supplementary Table 4). We required individuals to have at least 2 instances of a component code to be coded with any one of the compensation traits. In liver transplant cases, only ICD codes prior to the liver transplant CPT date were used. Decompensation counts were then determined by summing the number of coded decompensation traits for each individual (minimum=0, maximum=5) (Supplementary Table 5). Differences in average decompensation counts were assessed between males and females across the entire sample and within liver transplant, liver disease, and control definitions, using t-tests.

### Replication in All of Us

We replicated our findings in the All of Us Research Program, a population-based cohort that contains demographic, EHR, and survey information on participants. All of Us contains data on over 271,000 individuals of whom 56,715 had recorded lab values for component MELDNa labs and were included in the replication analyses (Supplementary Table 1). Even though the sample size of transplanted patients is small in All of Us (n=25), the broader sample allowed us to confirm the pattern of sex differences in MELDNa labs seen in VUMC and further demonstrate pervasive sex differences even in the absence of liver transplant or disease.

Definitions for controls, liver disease, liver transplant, and decompensation traits were the same as in VUMC. Lab values were filtered as described in VUMC and then used to calculate OPTN MELDNa scores (Supplementary Tables 5-6). Sex differences in median lab values, median calculated MELDNa scores, and decompensation counts were assessed across the entire sample and within liver status groups using t-tests.

### Development of Sex-Adjusted MELDNa Map

Individuals in VUMC with evidence of liver disease but who had not undergone a liver transplant were used to develop a sex-adjusted MELDNa score map. A fair MELDNa score should satisfy the property that a male and a female who have similar scores should also have similar liver disease severity. However, as shown in previous literature and corroborated by our work, current OPTN MELDNa score underestimates the degree of illness in females with liver disease. To reduce this disparity, our mapping focused on creation of a score such that mapped sex-adjusted scores resulted in more similar distributions of disease severity for males and females. We created the sex-adjusted MELDNa map with the following three steps. First, females and males were matched on race, age at MELDNa score, and decompensation count, which we used as the surrogate for the liver disease severity using nearest-neighbor matching in the MatchIt R package^20^. We divided the matched sample by sex and separately calculated quantiles of MELDNa scores. Because liver transplant waitlist listing dates are not available in EHRs, we used the first calculated MELDNa score after liver disease diagnosis to most closely estimate a patient’s listing MELDNa score on the transplant waitlist. Corresponding quantiles were matched to produce the final map. The quantile mapping reduces the underestimation bias of MELDNa scores for females and preserves the ranking of OPTN MELDNa scores within females. In other words, considering two females, the female that has a higher OPTN MELDNa score than the other would still have a larger (or equal) sex-adjusted MELD score. Additionally, this approach accommodates non-linearity in sex-differences which can result in a smaller sex difference in outcomes for the extreme high MELDNa scores.

### Evaluating Sex-Adjusted MELDNa Scores in LSAM

Because the sex-adjusted score was derived using EHR data which may include waitlisted and non-waitlisted patients, we next sought to validate the score in a liver transplant waitlist sample. Liver Simulated Allocation Modeling (LSAM) is a discrete event simulation that models the functioning of the US liver allocation system and allows for comparison of various allocation schemes^21^. The LSAM software was used to model outcomes of changing OPTN MELDNa scores to the proposed sex-adjusted scores. Simulations using both the OPTN MELDNa score and the proposed sex-adjusted MELDNa score were conducted with 10 replications of organ and waitlist arrivals from most recent liver transplant data (2015.6 –2016.6) contained in the LSAM package. Within each replication, the same simulated dataset was used for both MELDNa scores to generate comparative estimates of transplant volume, wait-list mortality, and the impact of these with regard to sex disparity in receiving liver transplant. The primary outcome of interest is the impact of the sex-adjusted MELDNa on sex-specific total transplant volume compared with the current MELDNa. Secondary outcomes included 1-year wait-list mortality (per 100 person-year) and total deaths.

### Statistical Analyses

All analyses were completed in R version 3.4 or 4.0.0 (R Foundation for Statistical Computing, Vienna, Austria).

## Results

### Sex differences in component labs and calculated MELDNa scores

Within controls, males had increased creatinine (*p*<2.22⨯10^−308^), INR (*p*=2.65⨯10^−114^), and bilirubin levels (*p*<2.22⨯10^−308^) compared to females, but sodium levels were not different (*p*=0.60). Males with liver disease showed increased creatinine (*p*=1.03⨯10^−291^), INR (*p*=1.80⨯10^−14^), and bilirubin levels (*p*=6.42⨯10^−54^), as well as decreased sodium levels compared to females (*p*=1.88×10^−23^). In liver transplant cases, males had increased pre-transplant creatinine levels compared to females (*p*=0.001). Pre-transplant INR (*p*=0.74), bilirubin (*p*=0.26), and sodium levels (*p*=0.36) were not significantly different between male and female transplant cases (Figure 1a-d, Supplementary Tables 7-8).

**Fig. 1.**
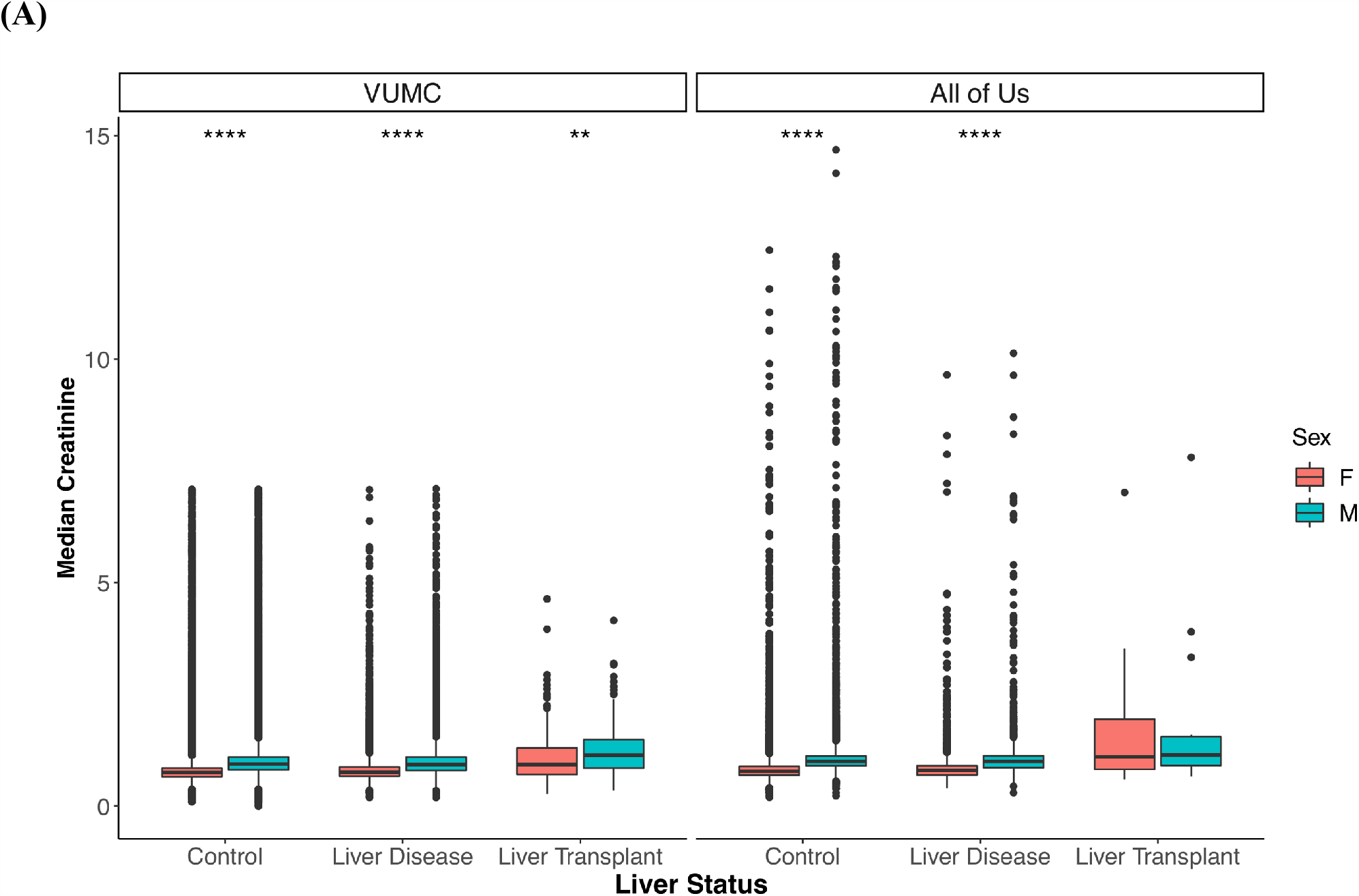
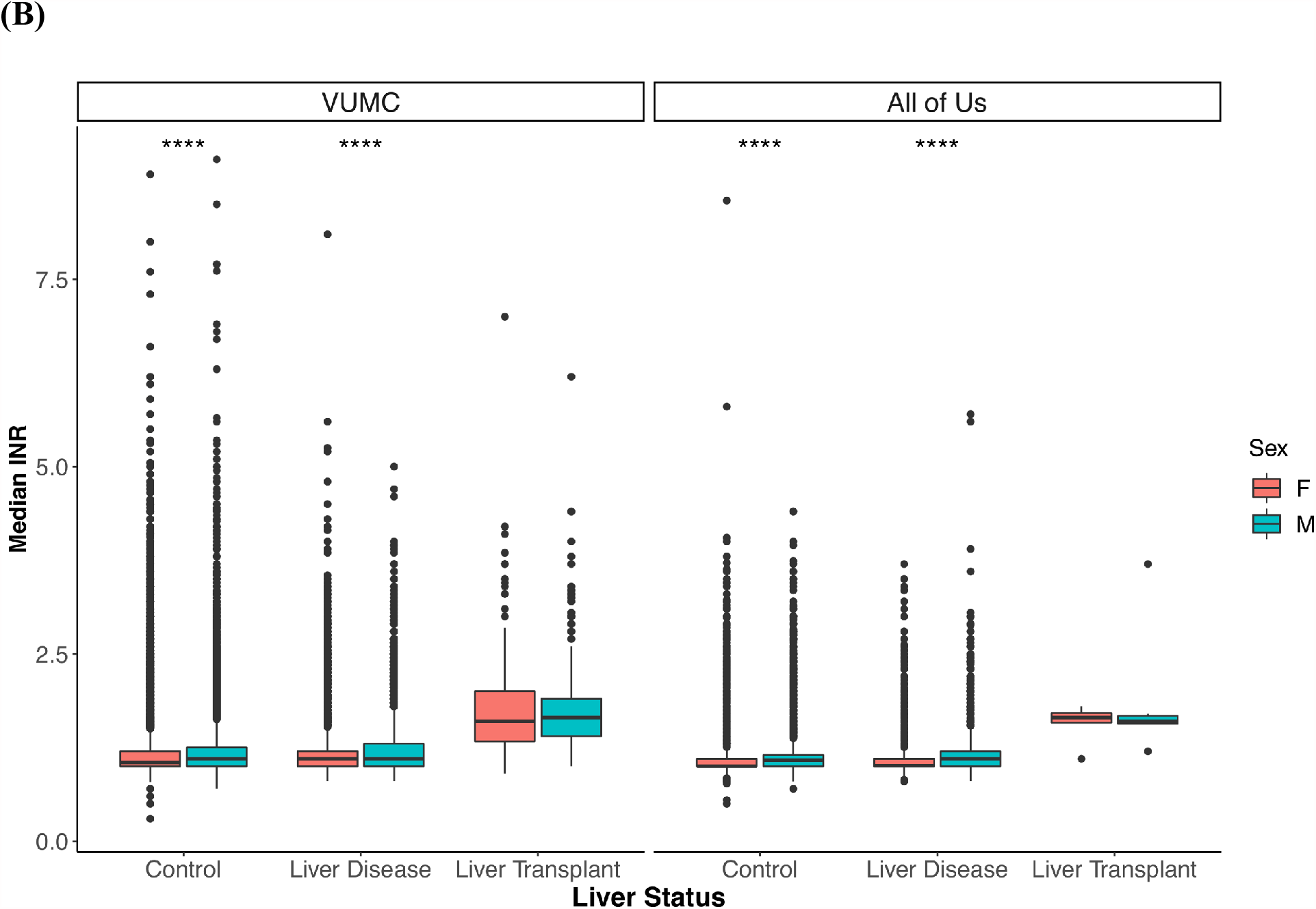
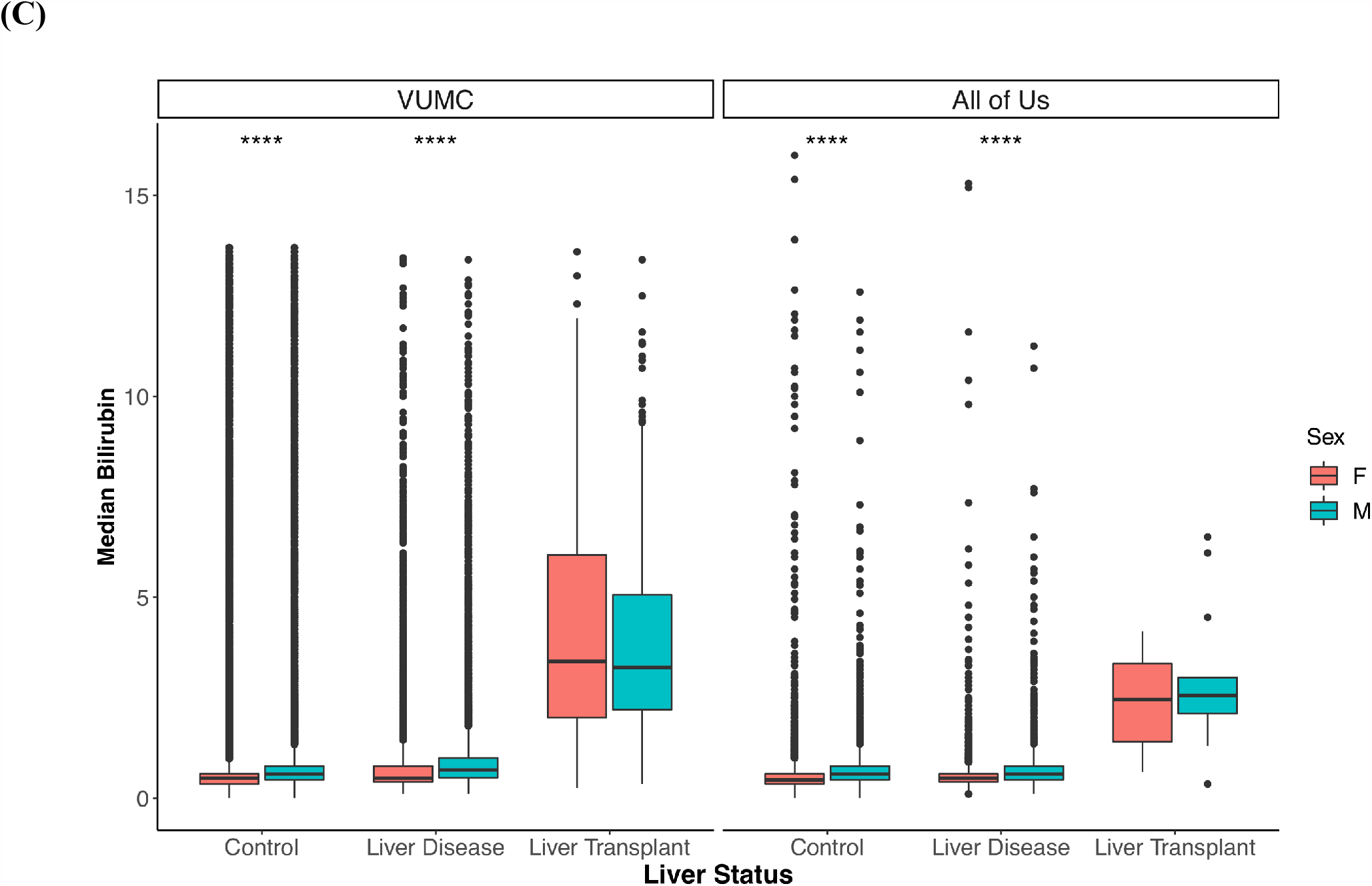
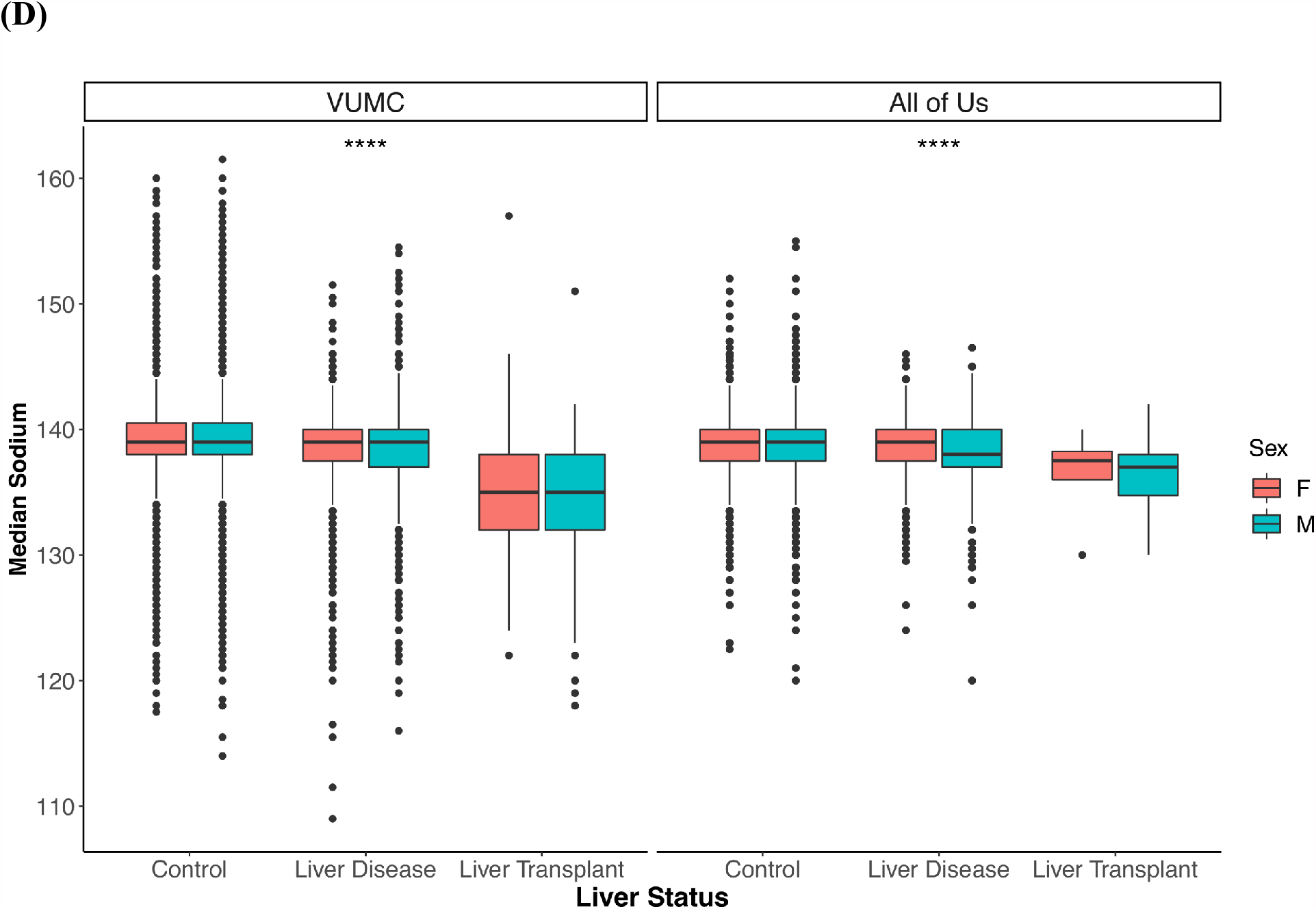
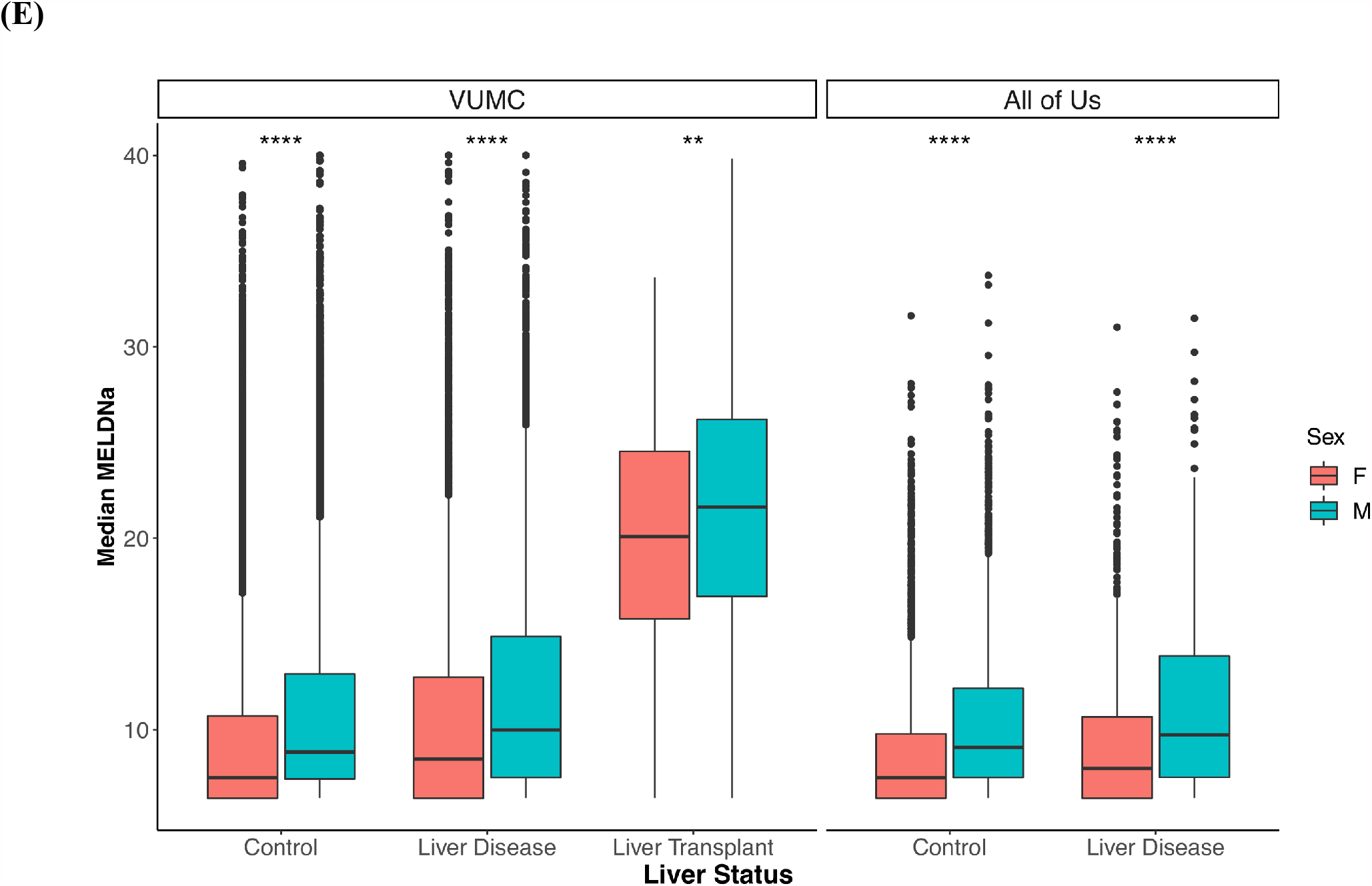
Sex differences in MELDNa lab values. Sex differences in median (A) creatinine, (B) INR, (C) bilirubin, (D) sodium levels, and (E) calculated OPTN MELDNa scores stratified by liver status in VUMC and All of Us. For liver transplant recipients, median levels prior to liver transplant date were utilized. For controls and liver disease cases, median levels across the entire EHR were used. Sex differences were assessed using a t-test.** denotes p<=0.01; ***: p<=0.001; ****: p<=0.0001.

Median MELDNa (MELDNa_median_) scores were increased in males compared to females within each liver status group (*p*_*controls*_=3.16×10^−316^, *p*_*liver disease*_ =2.20×10^−54^; *p*_*liver transplant*_*=*0.005) (Figure 1e, Supplementary Tables 7-8).

### Sex differences in decompensation counts

Among controls and liver disease cases, males had higher decompensation counts than females (*p*_*controls*_=2.86⨯10^−31^; *p*_*liver disease*_=2.75⨯10^−10^). However, among patients who had received a transplant, females had higher pre-transplant decompensation counts (*p*=0.005) (Figure 2, Supplementary Table 9). The difference in the average number of decompensation traits between liver disease cases and liver transplant cases was 1.4 for females and 1.1 for males, suggesting females generally accrue more decompensation traits prior to receiving a liver transplant.

**Fig. 2.**
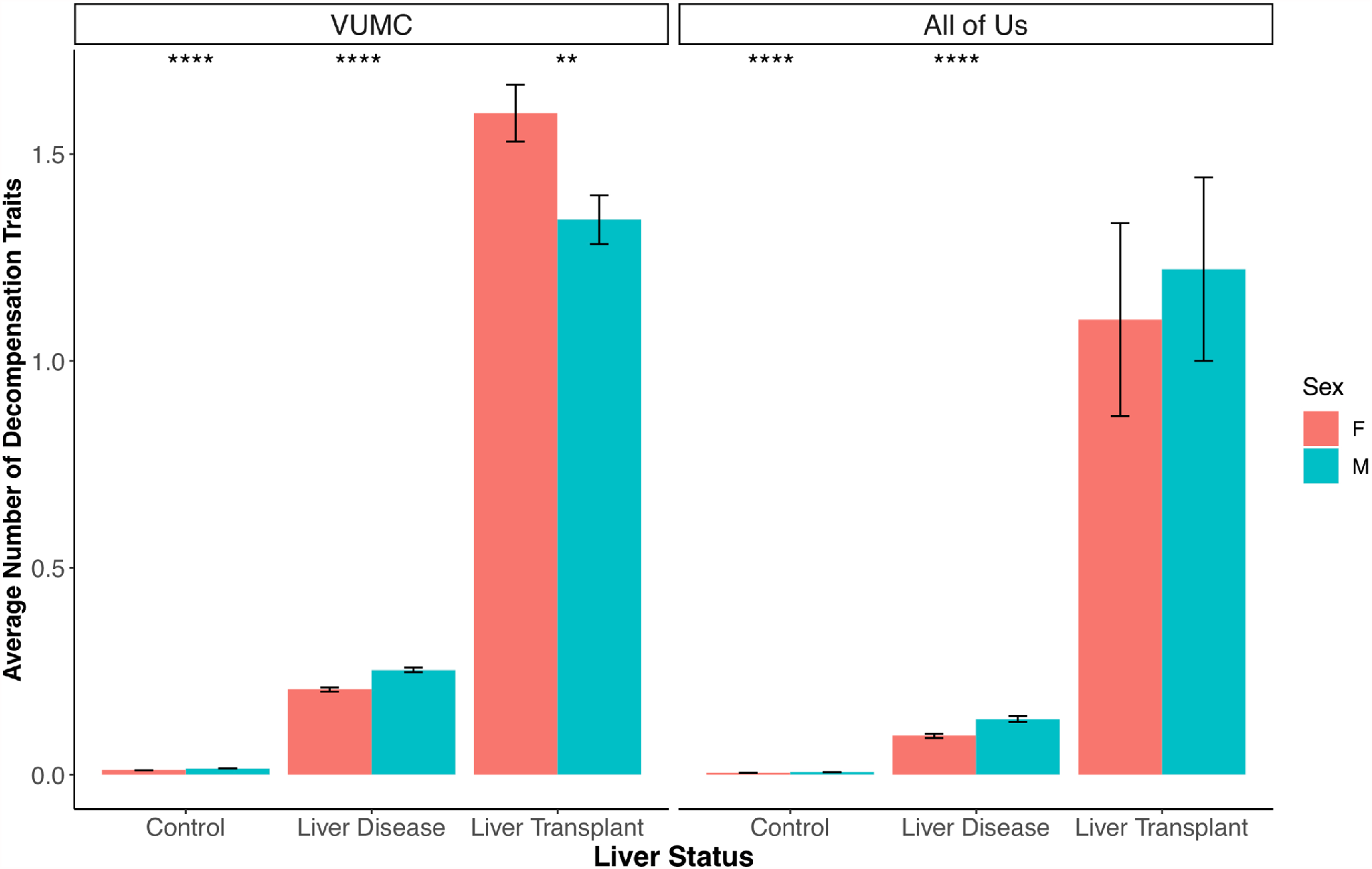
Sex differences in average number of decompensation traits stratified by liver status in VUMC and All of Us. For liver transplant recipients, decompensation traits prior to liver transplant date were utilized. For controls and liver disease cases, decompensation traits across the entire EHR were used. Sex differences were assessed using a t-test. ** denotes p<=0.01; ***: p<=0.001; ****: p<=0.0001.

**Fig. 3.**
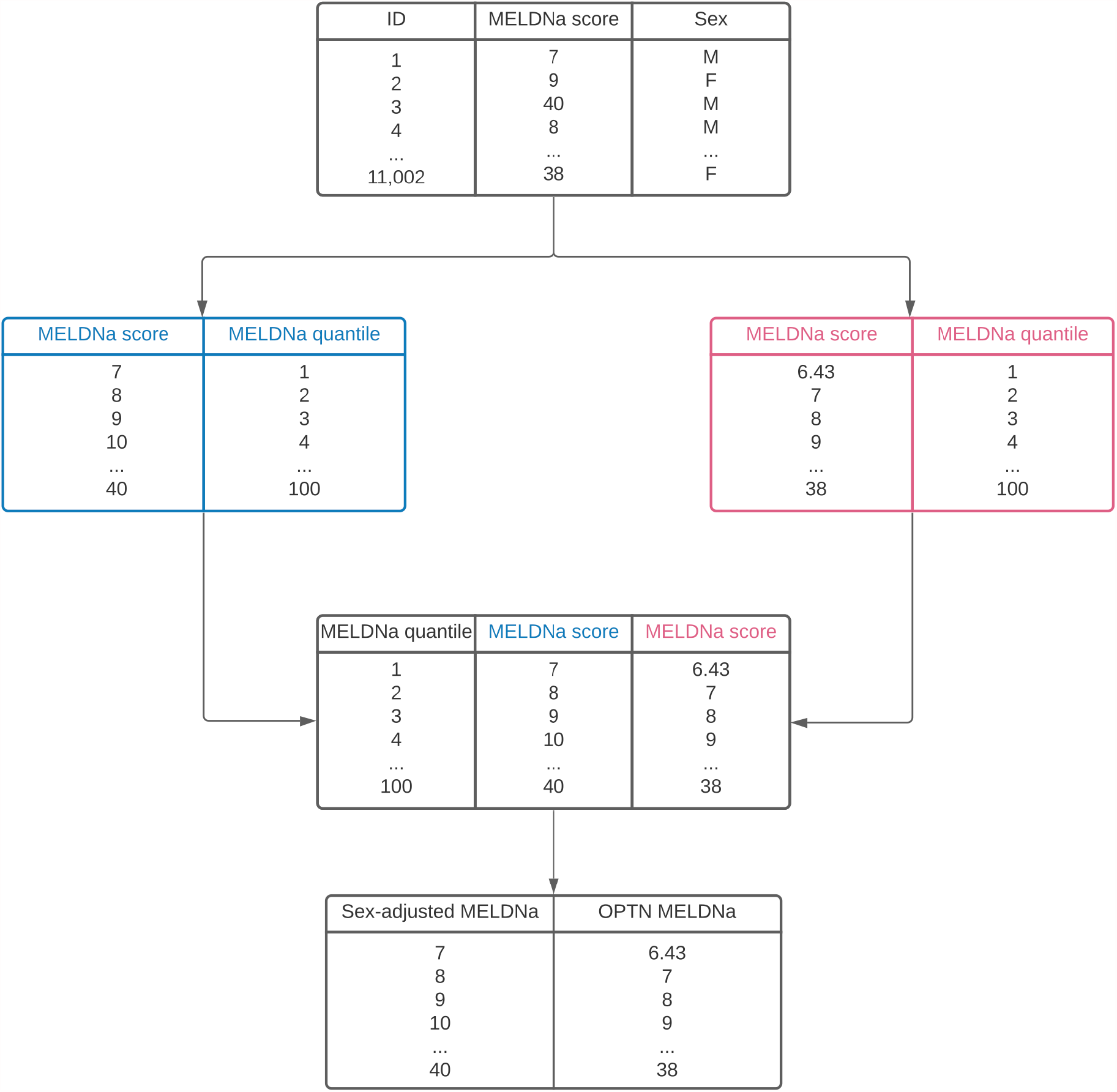
Derivation of the sex-adjusted MELDNa map. In VUMC, non-transplanted liver disease patients were matched by sex on number of decompensation traits, age at MELDNa score, and race. Next, quantiles of MELDNa scores were calculated separately for males and females. MELDNa scores were then matched based on quantile. The sex-adjusted map converts the original female OPTN score to the corresponding male score.

### Replication in All of Us

Across the entire All of Us sample, males had significantly higher median MELDNa lab values compared to females (*p*_creatinine_<2.23⨯10^−223^; *p*_*INR*_=8.48⨯10^−40^; *p*_bilirubin_=5.11⨯10^−169^; *p*_sodium_=4.08⨯10^−3^). In controls, males had higher median creatinine (*p*=1.22⨯10^−316^), INR (*p*=3.49⨯10^−31^), and bilirubin levels *(p*=4.44⨯10^−168^), but not sodium levels (p=0.643). Males had significantly higher median levels of all MELDNa labs within liver disease cases *(p*_creatinine_=8.39⨯10^−34^; *p*_INR_=1.43⨯10^−9^; *p*_bilirubin_=8.64⨯10^−13^; *p*_sodium_=3.16⨯10^−9^). Within liver transplant recipients, median lab values were not significantly different between males and females *(p*_creatinine_=0.96; *p*_INR_=0.53; *p*_bilirubin_=0.32; *p*_sodium_=0.57), however, it should be noted that sample size was small for this group (N<20; eTable 7, Supplementary Figure 3). In the comparison of median calculated MELDNa scores, males had significantly higher scores compared to females across the entire sample (*p*=3.31⨯10^−48^), controls (p=4.64⨯10^−33^), and liver disease cases (*p*=1.87⨯10^−14^).

Males had higher counts of decompensation traits across the entire sample (*p*=1.31⨯10^−16^), within controls (*p*=5.76⨯10^−6^), and within liver disease cases (*p*=3.15⨯10^−6^), but not within liver transplant recipients (p=0.71) (Supplementary Table 9, Supplementary Figure 4).

### Development of Sex-Adjusted MELDNa Map

Male MELDNa scores remain based on the OPTN calculation while the female sex-adjusted MELDNa score is found in the “sex-adjusted MELDNa” column corresponding to the unadjusted OPTN MELDNa score. Female scores were increased between 0-2 points and 1.6 points on average.

### Evaluating Sex-Adjusted MELDNa Scores in LSAM

Using the current OPTN MELDNa scores, males were transplanted 0.7% higher rate than females (males=23.7%; females=23.0%). When the sex-adjusted scores were assessed, females were transplanted at a 1% higher rate than males (males=23.1%; females=24.1%). Total death counts for both sexes decreased using the sex-adjusted scores and waitlist death rates decreased among females (OPTN=15%; sex-adjusted=14%) while remaining unchanged in males (both=16%) (Table 2).

**Table 1.** Proposed sex-adjusted MELDNa score map. Female sex-adjusted MELDNa scores are found in the “sex-adjusted MELDNa” column corresponding to the unadjusted OPTN MELDNa score. Male MELDNa scores remain based on the OPTN calculation.

| **Female OPTN MELDNa** | **Female Sex-Adjusted MELDNa** |
| --- | --- |
| 6 | 7 |
| 7 | 9 |
| 8 | 9 |
| 9 | 11 |
| 10 | 12 |
| 11 | 13 |
| 12 | 14 |
| 13 | 15 |
| 14 | 16 |
| 15 | 17 |
| 16 | 18 |
| 17 | 19 |
| 18 | 20 |
| 19 | 21 |
| 20 | 22 |
| 21 | 23 |
| 22 | 24 |
| 23 | 25 |
| 24 | 26 |
| 25 | 27 |
| 26 | 28 |
| 27 | 28 |
| 28 | 29 |
| 29 | 30 |
| 30 | 32 |
| 31 | 33 |
| 32 | 34 |
| 33 | 35 |
| 34 | 36 |
| 35 | 36 |
| 36 | 37 |
| 37 | 37 |
| 38 | 38 |
| 39 | 39 |
| 40 | 40 |

**Table 2.** Summary of LSAM simulation results comparing OPTN to sex-adjusted MELDNa scores. Results are averaged across 10 iterations using independent sets of organ and waitlist arrivals from June 2015 – June 2016.

| MELDNa Version | Sex | N participants | N transplants<br>(proportion) | N total 1-year death | N waitlist death<br>(proportion) |
| --- | --- | --- | --- | --- | --- |
| OPTN | All | 27,032 | 6,343 (23.5%) | 2,493 | 2,120 (16%) |
|  | Males | 16,855 | 3,999 (23.7%) | 1,613 | 1,373 (16%) |
|  | Females | 10,177 | 2,344 (23.0%) | 880 | 747 (15%) |
| Sex-adjusted | All | 27,032 | 6,345 (23.5%) | 2,480 | 2,114 (16%) |
|  | Males | 16,855 | 3,894 (23.1%) | 1,608 | 1,383 (16%) |
|  | Females | 10,177 | 2,451 (24.1%) | 872 | 731 (14%) |

## Discussion

The MELD allocation system is designed to treat each individual equally, however, equality does not always ensure equity. Uncharacterized differences between groups in clinical lab values can exacerbate health disparities. Previous studies ascribed sex differences in MELDNa scores to known sex differences in creatinine levels^3,16–18^, but creatinine does not fully account for the sex difference in MELDNa scores^3,11,18^. Using two EHR systems, we analyzed sex differences in all component labs of MELDNa at scale and in diverse clinical presentations (i.e. controls, liver disease, and transplanted individuals). Our analyses showed significant sex differences in all four component labs of MELDNa, which systematically disadvantaged females. Even though the differences were small, the use of a natural logarithm of each lab value in the MELDNa calculation magnifies the differences, resulting in increased baseline MELDNa scores in males versus females.

Females are more likely to die or be de-listed due to disease severity while on the liver transplant waitlist^8,10,12^. Concordantly, in our sample, females who had undergone a liver transplant had significantly more pre-transplant decompensation traits than males. However, among liver disease cases who had not undergone a transplant, females had fewer decompensation traits. This pattern of results suggests that by the time females qualify for liver transplant, they are, on average, sicker than males, consistent with previous reports of higher waitlist mortality or removal for females^3,4,8,10^. Furthermore, female liver transplant recipients had lower MELDNa scores despite having more decompensation traits than male transplant recipients, demonstrating that OPTN MELDNa scores may not accurately reflect liver disease progression in females. Although not examined in this study, previous studies confirmed height and liver size contribute to the sex disparity in liver transplant. However, even after controlling for height or estimated liver size, rates of transplant remained lower among females^4,14,22^

The use of EHRs provides a unique view of sex difference in MELDNa scoring. Whereas previous studies were limited to only individuals with advanced liver disease, we were able to examine sex differences across multiple clinical endpoints. Additionally, we investigated sex differences in all MELDNa component labs, rather than restricting to only creatinine. Finally, the use of ICD codes enabled us to determine degree of liver decompensation to provide another metric of disease severity apart from MELDNa scoring.

We proposed and tested a sex-adjusted MELDNa score map to compensate for the sex differences seen in MELDNa component labs. The sex-adjusted score empirically matches a female’s MELDNa score to a male’s score of equal illness. The majority of sex-adjusted scores for females increase 1 or 2 points from the corresponding OPTN MELDNa score, consistent with a previous report of increased liver transplant access for females with an additional 2 points added to their MELD scores^15^. While a blanket addition of 1-2 points may over-correct female scores, especially at higher scores, our proposed adjustment ameliorates this issue by providing a score anchored to disease severity (i.e. decompensation trait count). The LSAM models using the sex-adjusted scores showed an increase in female transplantation rate and a decrease in overall death for both sexes, suggesting modest but positive improvement in reducing the sex disparity. Using the sex-adjusted scores, females had a 1% higher transplantation rate (compared to a 0.7% higher rate for males using the OPTN scores). While this adjustment resulted in a slight increase in female transplant rate compared to males, it is not possible to determine if this is due to an over-correction for female scores or if females on the waitlist had more severe disease that warranted a higher transplantation rate. Regardless, total death counts were reduced for both males and females and reduced female waitlist mortality, suggested that the sex-adjusted scores could help save lives.

### Limitations

Analysis of EHR data comes with several limitations. EHRs are not linked to national databases, such as the liver transplant waitlist, preventing us from integrating listing and de-listing dates into our analyses. Additionally, clinical lab tests are not ordered uniformly, potentially creating diagnostic bias in our sample, especially in controls without liver disease.

The proposed sex-adjusted MELDNa score does not solve other known causes of sex disparities including donor-recipient size mismatch or geographic or racial disparities. To successfully eliminate the sex disparity in liver transplant, further investigation and potential policy changes, such as access to pediatric donors for female recipients, increase in utilization of partial liver transplant for female recipients, and increase sharing of donors across UNOS regions, are necessary.

### Conclusions

Using EHR data, we demonstrate all lab traits used in the calculation of MELDNa scores show sex differences that increase males scores compared to females, despite females showing greater liver decompensation. In simulations, our proposed sex-adjusted MELDNa score increases the rate of female transplantation and decreases overall death in both sexes.

## Data Availability

Summary level data is available in the Supplementary Information.

## Abbreviations

MELD: model for end-stage liver disease
INR: international normalized ratio
OPTN: Organ Procurement and Transplantation Network
eGFR: estimated glomerular filtration rate
VUMC: Vanderbilt University Medical Center
EHR: electronic health record
ICD: International Classification of Disease
CPT: Current Procedural Terminology
UNOS: United Network for Organ Sharing
LSAM: Liver Simulated Allocation Modeling

## Acknowledgements

Dr. Guanhua Chen and Dr. Lea Davis are co-corresponding authors of the work.

CTSA (SD, Vanderbilt Resources)

The project described was supported by the National Center for Research Resources, Grant UL1 RR024975-01, and is now at the National Center for Advancing Translational Sciences, Grant 2 UL1 TR000445-06. The content is solely the responsibility of the authors and does not necessarily represent the official views of the NIH.

The *All of Us* Research Program is supported by the National Institutes of Health, Office of the Director: Regional Medical Centers: 1 OT2 OD026549; 1 OT2 OD026554; 1 OT2 OD026557; 1 OT2 OD026556; 1 OT2 OD026550; 1 OT2 OD 026552; 1 OT2 OD026553; 1 OT2 OD026548; 1 OT2 OD026551; 1 OT2 OD026555; IAA #: AOD 16037; Federally Qualified Health Centers: HHSN 263201600085U; Data and Research Center: 5 U2C OD023196; Biobank: 1 U24 OD023121; The Participant Center: U24 OD023176; Participant Technology Systems Center: 1 U24 OD023163; Communications and Engagement: 3 OT2 OD023205; 3 OT2 OD023206; and Community Partners: 1 OT2 OD025277; 3 OT2 OD025315; 1 OT2 OD025337; 1 OT2 OD025276. In addition, the All of Us Research Program would not be possible without the partnership of its participants.

## Citations

1. Wiesner, R. et al. Model for end-stage liver disease (MELD) and allocation of donor livers. Gastroenterology 124, 91–96 (2003).

2. Bernardi, M., Gitto, S. & Biselli, M. The MELD score in patients awaiting liver transplant: Strengths and weaknesses. J. Hepatol. 54, 1297–1306 (2011).

3. Myers, R. P., Shaheen, A. A. M., Aspinall, A. I., Quinn, R. R. & Burak, K. W. Gender, renal function, and outcomes on the liver transplant waiting list: Assessment of revised MELD including estimated glomerular filtration rate. J. Hepatol. 54, 462–470 (2011).

4. Lai, J. C., Terrault, N. A., Vittinghoff, E. & Biggins, S. W. Height contributes to the gender difference in wait-list mortality under the MELD-based liver allocation system. Am. J. Transplant. 10, 2659–2664 (2010).

5. Kim, W. R. et al. Hyponatremia and mortality among patients on the liver-transplant waiting list. N. Engl. J. Med. 359, 1018–1026 (2008).

6. Ruf, A. E. et al. Addition of serum sodium into the MELD score predicts waiting list mortality better than MELD alone. Liver Transplant. 11, 336–343 (2005).

7. Mathur, A. K., Schaubel, D. E., Gong, Q., Guidinger, M. K. & Merion, R. M. Sex-based disparities in liver transplant rates in the United States. Am. J. Transplant. 11, 1435–1443 (2011).

8. Moylan, C. a et al. Disparities in Liver Transplantation Before and After Introduction of the MELD Score. Jama 300, 2371–2378 (2013).

9. Bryce, C. L. et al. Sociodemographic differences in early access to liver transplantation services. Am. J. Transplant. 9, 2092–2101 (2009).

10. Cullaro, G., Sarkar, M. & Lai, J. C. Sex-based disparities in delisting for being “too sick” for liver transplantation. Am. J. Transplant. 18, 1214–1219 (2018).

11. Sarkar, M., Watt, K. D., Terrault, N. & Berenguer, M. Outcomes in liver transplantation: Does sex matter? J. Hepatol. 62, 946–955 (2015).

12. Locke, J. E. et al. Quantifying Sex-Based Disparities in Liver Allocation. JAMA Surg. (2020). doi:10.1001/jamasurg.2020.1129

13. Nephew, L. D. et al. Exception Points and Body Size Contribute to Gender Disparity in Liver Transplantation. Clin. Gastroenterol. Hepatol. 15, 1286-1293.e2 (2017).

14. Mindikoglu, A. L., Emre, S. H. & Magder, L. S. Impact of Estimated Liver Volume and Liver Weight on Gender Disparity in Liver Transplantation. Liver Transplant. 13, 767– 768 (2007).

15. Allen, A. M. et al. Reduced Access to Liver Transplantation in Women: Role of Height, MELD Exception Scores, and Renal Function Underestimation. Transplantation 102, 1710–1716 (2018).

16. Mindikoglu, A. L., Regev, A., Seliger, S. L. & Magder, L. S. Gender Disparity in Liver Transplant Waiting-List Mortality: The Importance of Kidney Function. Liver Transplant. 16, 1147–2257 (2010).

17. Cholongitas, E. et al. Female liver transplant recipients with the same GFR as male recipients have lower MELD scores - A systematic bias. Am. J. Transplant. 7, 685–692 (2007).

18. Huo, S. C. et al. Is the corrected-creatinine model for end-stage liver disease a feasible strategy to adjust gender difference in organ allocation for liver transplantation? Transplantation 84, 1406–1412 (2007).

19. Denny, J. C. et al. The ‘all of us’ research program. N. Engl. J. Med. (2019). doi:10.1056/NEJMsr1809937

20. Ho, D. E., Imai, K., King, G. & Stuart, E. A. MatchIt: Nonparametric preprocessing for parametric causal inference. J. Stat. Softw. (2011). doi:10.18637/jss.v042.i08

21. Thompson, D. et al. Simulating the allocation of organs for transplantation. Health Care Manag. Sci. 7, 331–338 (2004).

22. Darden, M., Parker, G., Anderson, E. & Buell, J. F. Persistent sex disparity in liver transplantation rates. Surg. (United States) 169, 694–699 (2021).

