## Supplementary Material for "Making an equal system equitable: Proposing a sex-adjusted MELDNa score for liver transplantation allocation"

*Supplementary Figure 2. Decompensation counts by median calculated MELDNa scores stratified by sex and liver disease status…………………………………………………………6*

### *Calculation of OPTN MELDNa Scores*

Calculation of OPTN MELDNa scores followed current clinical guidelines. Creatinine, INR, and bilirubin values were set to a minimum of 1 and creatinine levels were set to a maximum of 4. Sodium levels were restricted to a minimum of 125 and a maximum of 137. Calculated OPTN MELDNa scores were restricted to a maximum of 40. OPTN MELDNa was then calculated using the OPTN formula^20^:

*MELD = 3.78*ln(bilirubin) + 11.2*ln(INR) + 9.57*ln(creatinine) + 6.43*

*MELDNa = MELD + 1.32*(137 – sodium) – [0.033*MELD*(137 – sodium)]*

### *Decompensation Count by MELDNa Score*

To determine how OPTN MELDNa scores track with the number of decompensation traits among males and females, linear regression models were fit between OPTN MELDNa_median_ scores and decompensation counts. Analyses were stratified by liver status groups and sex (Supplementary Figure 2).

### *Replication in All of Us Research Program*

In All of Us controls, males had higher median creatinine (*p*=1.22x10^-316^), INR (*p*=3.49x10^-31^), and bilirubin levels *(p*=4.44x10^-168^)*,* but not sodium levels (p=0.643). Males had significantly higher median levels of all MELDNa labs within liver disease cases (*p_creatinine_*=8.39x10^-34^; *p_INR_*=1.43x10^-9^; *p_bilirubin_*=8.64x10^-13^; *p_sodium_*=3.16x10^-9^)*.* Within liver transplant recipients, median lab values were not significantly different between males and females (*p_creatinine_*=0.96; *p_INR_*=0.53; *p_bilirubin_*=0.32; *p_sodium_*=0.57), however, it should be noted that sample size was small for this group (N<20; eTable 7, eFigure 3). In the comparison of median calculated MELDNa scores, males had significantly higher scores compared to females in controls (*p*=4.64x10^-33^) and liver disease cases (*p*=1.87x10^-14^).

Males had higher counts of decompensation traits within controls (*p*=5.76x10^-6^) and within liver disease cases (*p*=3.15x10^-6^), but not within liver transplant recipients (*p*=0.71) (eTable 9, eFigure 4).

Supplementary Figure 1. Sex differences in maximum a) creatinine, b) INR, c) bilirubin, d) sodium, and e) calculated MELDNa stratified by liver status in VUMC. In liver disease cases and controls, maximum values across the entire medical record were selected. In liver transplant cases, maximum before transplant was selected. Statistical significance was determined with Student’s t-tests.

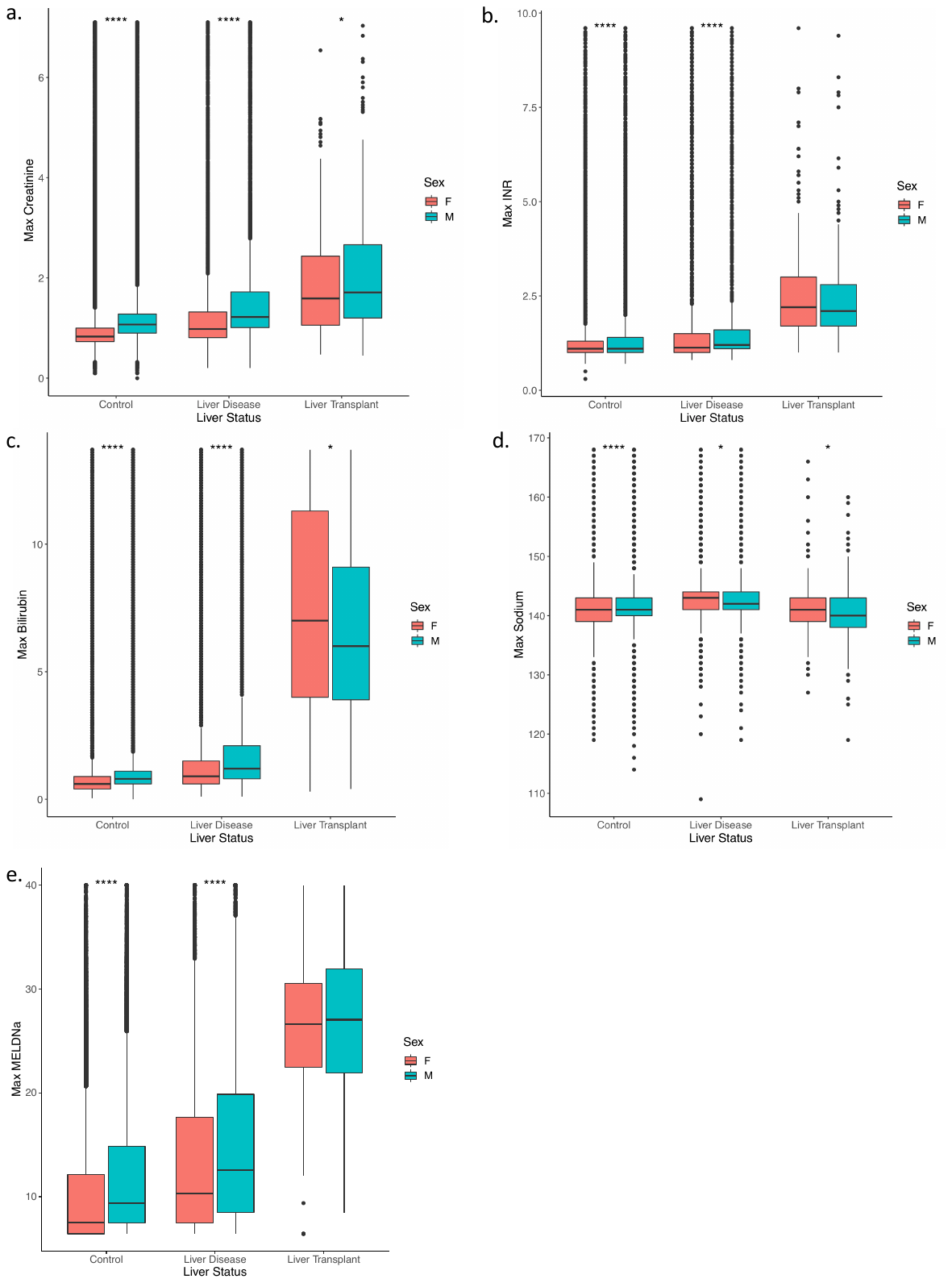
Supplementary Figure 2. Decompensation counts by median calculated MELDNa stratified by sex and liver status. A linear regression was fit between the number of decompensation traits and median MELDNa scores.

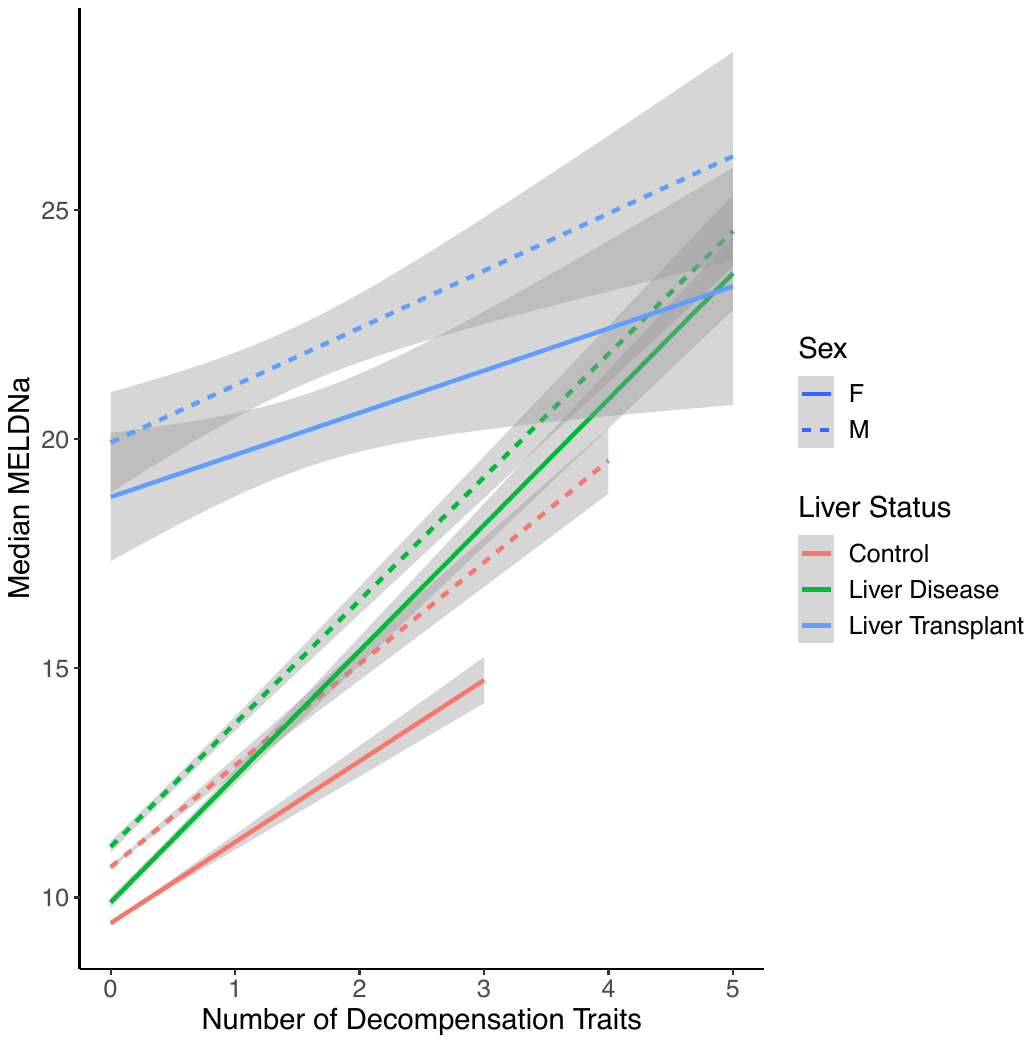

### Supplementary Table 1. Sample characteristics of Vanderbilt University Medical Center Synthetic Derivative (VUMC) and All of Us.

|  | **Sample Source** | **All** | **Males** | **Females** |
| --- | --- | --- | --- | --- |
| **N** | VUMC | 623,931 | 263,955 | 359,976 |
|  | All of Us | 56,715 | 19,715 | 36,154 |
| **Liver Transplant Cases** | VUMC | 601 | 354 | 247 |
|  | All of Us | 25 | <=20 | <=20 |
| **Liver Disease Cases** | VUMC | 24,921 | 12,247 | 12,674 |
|  | All of Us | 6,422 | 2,756 | 3,666 |
| **Controls** | VUMC | 598,409 | 251,354 | 347,055 |
|  | All of Us | 49,422 | 16,944 | 32,478 |
| **Median age (IQR)** | VUMC | 44 (23 – 61) | 45 (19 – 62) | 43 (25 – 60) |
|  | All of Us | 58 (43 - 68) | 60 (47 - 69) | 57 (42 - 67) |
| **% White** | VUMC | 81.40% | 82.50% | 80.60% |
|  | All of Us | 53.65% | 55.73% | 52.85% |

Supplementary Table 3. Descriptive statistics of median MELDNa component labs and median calculated MELDNa stratified by sex and liver status in VUMC. For liver disease cases and controls, median was calculated across the entire medical record. In liver transplant cases, median was calculated before the liver transplant date.

| **Lab** | **Liver Status** | **Sex** | **N** | **Median** | **Mean** | **SD** | **Range** | **IQR** |
| --- | --- | --- | --- | --- | --- | --- | --- | --- |
| Creatinine | All | All | 414,374 | 0.82 | 0.88 | 0.36 | 0 - 7.1 | 0.7 - 0.98 |
|  |  | Female | 237,338 | 0.75 | 0.79 | 0.3 | 0.1 - 7.09 | 0.66 - 0.85 |
|  |  | Male | 177,036 | 0.94 | 0.99 | 0.39 | 0 - 7.1 | 0.81 - 1.1 |
|  | Control | All | 389,459 | 0.81 | 0.87 | 0.35 | 0 - 7.09 | 0.7 - 0.98 |
|  |  | Female | 224,706 | 0.75 | 0.79 | 0.29 | 0.1 - 7.09 | 0.66 - 0.85 |
|  |  | Male | 164,753 | 0.94 | 0.99 | 0.39 | 0 - 7.09 | 0.81 - 1.1 |
|  | Liver Disease | All | 24,339 | 0.83 | 0.92 | 0.43 | 0.2 - 7.1 | 0.71 - 1 |
|  |  | Female | 12,385 | 0.76 | 0.83 | 0.36 | 0.2 - 7.08 | 0.67 - 0.88 |
|  |  | Male | 11,954 | 0.93 | 1.02 | 0.46 | 0.2 - 7.1 | 0.8 - 1.1 |
|  | Liver Transplant | All | 576 | 1.06 | 1.19 | 0.56 | 0.27 - 4.63 | 0.8 - 1.42 |
|  |  | Female | 247 | 0.93 | 1.11 | 0.57 | 0.27 - 4.63 | 0.71 - 1.3 |
|  |  | Male | 329 | 1.14 | 1.26 | 0.55 | 0.35 - 4.16 | 0.85 - 1.49 |
| INR | All | All | 163,161 | 1.1 | 1.22 | 0.41 | 0.3 - 9.1 | 1 - 1.2 |
|  |  | Female | 84,502 | 1.1 | 1.20 | 0.4 | 0.3 - 8.9 | 1 - 1.2 |
|  |  | Male | 78,659 | 1.1 | 1.24 | 0.42 | 0.7 - 9.1 | 1 - 1.25 |
|  | Control | All | 142,701 | 1.1 | 1.22 | 0.41 | 0.3 - 9.1 | 1 - 1.2 |
|  |  | Female | 74,404 | 1.05 | 1.19 | 0.4 | 0.3 - 8.9 | 1 - 1.2 |
|  |  | Male | 68,297 | 1.1 | 1.24 | 0.42 | 0.7 - 9.1 | 1 - 1.25 |
|  | Liver Disease | All | 19,884 | 1.1 | 1.22 | 0.37 | 0.8 - 8.1 | 1 - 1.25 |
|  |  | Female | 9,851 | 1.1 | 1.20 | 0.36 | 0.8 - 8.1 | 1 - 1.2 |
|  |  | Male | 10,033 | 1.1 | 1.24 | 0.37 | 0.8 - 5 | 1 - 1.3 |
|  | Liver Transplant | All | 576 | 1.6 | 1.79 | 0.65 | 0.9 - 7 | 1.4 - 1.95 |
|  |  | Female | 247 | 1.6 | 1.80 | 0.73 | 0.9 - 7 | 1.34 - 2 |
|  |  | Male | 329 | 1.65 | 1.78 | 0.59 | 1 - 6.2 | 1.4 - 1.9 |
| Bilirubin | All | All | 338,641 | 0.5 | 0.65 | 0.73 | 0 - 13.7 | 0.4 - 0.7 |
|  |  | Female | 194,778 | 0.5 | 0.58 | 0.64 | 0 - 13.7 | 0.35 - 0.62 |
|  |  | Male | 143,863 | 0.6 | 0.76 | 0.83 | 0 - 13.7 | 0.45 - 0.8 |
|  | Control | All | 313,985 | 0.5 | 0.63 | 0.67 | 0 - 13.7 | 0.4 - 0.7 |
|  |  | Female | 182,258 | 0.5 | 0.56 | 0.59 | 0 - 13.7 | 0.35 - 0.6 |
|  |  | Male | 131,727 | 0.6 | 0.73 | 0.77 | 0 - 13.7 | 0.45 - 0.8 |
|  | Liver Disease | All | 24,116 | 0.6 | 0.88 | 1.06 | 0.1 - 13.45 | 0.4 - 0.9 |
|  |  | Female | 12,295 | 0.5 | 0.78 | 1.00 | 0.1 - 13.45 | 0.4 - 0.8 |
|  |  | Male | 11,821 | 0.7 | 0.99 | 1.11 | 0.1 - 13.4 | 0.5 - 1 |
|  | Liver Transplant | All | 540 | 3.3 | 4.11 | 2.76 | 0.25 - 13.6 | 2.1 - 5.32 |
| **Lab** | **Liver Status** | **Sex** | **N** | **Median** | **Mean** | **SD** | **Range** | **IQR** |
|  |  | Female | 225 | 3.40 | 4.27 | 3.01 | 0.25 - 13.6 | 2 - 6.05 |
|  |  | Male | 315 | 3.25 | 4.00 | 2.57 | 0.35 - 13.4 | 2.2 - 5.05 |
| Sodium | All | All | 408,746 | 139.00 | 139.02 | 2.31 | 109 - 161.5 | 138 - 140.5 |
|  |  | Female | 232,661 | 139.00 | 139.03 | 2.28 | 109 - 160 | 138 - 140 |
|  |  | Male | 176,085 | 139.00 | 139.00 | 2.36 | 114 - 161.5 | 138 - 140.5 |
|  | Control | All | 383,855 | 139.00 | 139.06 | 2.29 | 114 - 161.5 | 138 - 140.5 |
|  |  | Female | 220,055 | 139.00 | 139.06 | 2.26 | 117.5 - 160 | 138 - 140.5 |
|  |  | Male | 163,800 | 139.00 | 139.06 | 2.32 | 114 - 161.5 | 138 - 140.5 |
|  | Liver Disease | All | 24,315 | 139.00 | 138.5.0 | 2.47 | 109 - 154.5 | 137 - 140 |
|  |  | Female | 12,359 | 139.00 | 138.65 | 2.39 | 109 - 151.5 | 137.5 - 140 |
|  |  | Male | 11,956 | 139.00 | 138.34 | 2.54 | 116 - 154.5 | 137 - 140 |
|  | Liver Transplant | All | 576 | 135.00 | 134.60 | 4.54 | 118 - 157 | 132 - 138 |
|  |  | Female | 247 | 135.00 | 134.80 | 4.34 | 122 - 157 | 132 - 138 |
|  |  | Male | 329 | 135.00 | 134.46 | 4.69 | 118 - 151 | 132 - 138 |
| MELDNa | All | All | 98,015 | 8.47 | 10.43 | 4.99 | 6.43 - 40 | 6.66 - 12.37 |
|  |  | Female | 49,795 | 7.5.0 | 9.78 | 4.70 | 6.43 - 40 | 6.43 - 11.11 |
|  |  | Male | 48,220 | 9.07 | 11.09 | 5.19 | 6.43 - 40 | 7.5 - 13.41 |
|  | Control | All | 80,354 | 8.21 | 10.16 | 4.73 | 6.43 - 40 | 6.43 - 11.85 |
|  |  | Female | 41,152 | 7.50 | 9.54 | 4.44 | 6.43 - 39.58 | 6.43 - 10.72 |
|  |  | Male | 39,202 | 8.83 | 10.8 | 4.93 | 6.43 - 40 | 7.43 - 12.9 |
|  | Liver Disease | All | 17,122 | 9.27 | 11.36 | 5.63 | 6.43 - 40 | 7.43 - 13.91 |
|  |  | Female | 8,418 | 8.47 | 10.68 | 5.40 | 6.43 - 40 | 6.43 - 12.75 |
|  |  | Male | 8,704 | 9.98 | 12.01 | 5.77 | 6.43 - 40 | 7.5 - 14.86 |
|  | Liver Transplant | All | 539 | 20.86 | 21.09 | 6.17 | 6.43 - 39.82 | 16.35 - 25.62 |
|  |  | Female | 225 | 20.09 | 20.21 | 6.15 | 6.43 - 33.63 | 15.8 - 24.54 |
|  |  | Male | 314 | 21.62 | 21.72 | 6.11 | 6.43 - 39.82 | 16.96 - 26.2 |

### Supplementary Table 4. Decompensation phenotype ICD9 and ICD10 codes.

| ICD code | Description |
| --- | --- |
| 578.0 | Hematemesis |
| 578.9 | Hemorrhage of gastrointestinal tract, unspecified |
| 789.5 | Ascites |
| 789.51 | Malignant ascites |
| 789.59 | Other ascites |
| 782.4 | Jaundice, unspecified, not of newborn |
| 572.2 | Hepatic encephalopathy |
| K92.0 | Hematemesis |
| K92.2 | Gastrointestinal hemorrhage, unspecified |
| K70.11 | Alcoholic hepatitis with ascites |
| K70.31 | Alcoholic cirrhosis of liver with ascites |
| R17 | Unspecified jaundice |
| K72.9 | Hepatic failure, unspecified |
| K72.90 | Hepatic failure, unspecified without coma |
| K72.91 | Hepatic failure, unspecified with coma |

### Supplementary Table 5. Number of individuals with decompensation traits stratified by sex and liver status.

|  | **Sample** | **N decompensated (%)** | **N males decompensated (%)** | **N females decompensated (%)** |
| --- | --- | --- | --- | --- |
| **All** | VUMC | 12,297 (1.97%) | 6,284 (2.38%) | 6,013 (1.67%) |
|  | All of Us | 249 (0.11%) | 140 (0.17%) | 109 (0.08%) |
| **Controls** | VUMC | 7,346 (1.23%) | 3,602 (1.43%) | 3,744 (1.08%) |
|  | All of Us | 43 (0.02%) | 22 (0.03%) | 21 (0.02%) |
| **Liver Disease** | VUMC | 4,465 (17.91%) | 2,409 (19.67%) | 2,056 (16.22%) |
|  | All of Us | 195 (2.09%) | 110 (2.77%) | 85 (1.59%) |
| **Liver Transplant** | VUMC | 486 (80.87%) | 273 (77.12%) | 213 (86.23%) |
|  | All of Us | <=20 (39.3%) | <=20 (44.44%) | <=20 (30.0%) |

### Supplementary Table 6. Descriptive statistics of MELDNa component labs in the All of Us Research Program.

| **Lab** | **Liver Status** | **Sex** | **N** | **Median** | **Mean** | **SD** | **Range** | **IQR** |
| --- | --- | --- | --- | --- | --- | --- | --- | --- |
| Creatinine | All | All | 37,962 | 0.82 | 0.92 | 0.62 | 0.2 - 14.69 | 0.7 - 1 |
|  |  | Female | 25,529 | 0.78 | 0.83 | 0.45 | 0.2 - 12.44 | 0.7 - 0.89 |
|  |  | Male | 12,433 | 1 | 1.13 | 0.84 | 0.23 - 14.69 | 0.89 - 1.13 |
|  | Control | All | 32,099 | 0.81 | 0.90 | 0.54 | 0.2 - 14.69 | 0.7 - 0.99 |
|  |  | Female | 21,929 | 0.78 | 0.81 | 0.40 | 0.2 - 12.44 | 0.69 - 0.88 |
|  |  | Male | 10,170 | 1 | 1.10 | 0.73 | 0.23 - 14.69 | 0.9 - 1.12 |
|  | Liver Disease | All | 5,803 | 0.85 | 1.03 | 0.91 | 0.3 - 12.66 | 0.72 - 1 |
|  |  | Female | 3,579 | 0.8 | 0.89 | 0.64 | 0.4 - 10.8 | 0.7 - 0.9 |
|  |  | Male | 2,224 | 1 | 1.25 | 1.20 | 0.3 - 12.66 | 0.86 - 1.2 |
|  | Liver Transplant | All | 60 | 1.01 | 1.57 | 1.54 | 0.5 - 7.8 | 0.85 - 1.4 |
|  |  | Female | 21 | 0.94 | 1.48 | 1.46 | 0.5 - 7.02 | 0.73 - 1.5 |
|  |  | Male | 39 | 1.02 | 1.62 | 1.59 | 0.6 - 7.8 | 0.88 - 1.36 |
| INR | All | All | 23,495 | 1.04 | 1.13 | 0.32 | 0.5 - 8.55 | 1 - 1.1 |
|  |  | Female | 14,083 | 1 | 1.11 | 0.30 | 0.5 - 8.55 | 1 - 1.1 |
|  |  | Male | 9,412 | 1.1 | 1.17 | 0.34 | 0.7 - 5.7 | 1 - 1.2 |
|  | Control | All | 17,274 | 1.02 | 1.12 | 0.31 | 0.5 - 8.55 | 1 - 1.1 |
|  |  | Female | 10,578 | 1 | 1.10 | 0.30 | 0.5 - 8.55 | 1 - 1.1 |
|  |  | Male | 6,696 | 1.07 | 1.15 | 0.33 | 0.7 - 4.4 | 1 - 1.15 |
|  | Liver Disease | All | 6,197 | 1.05 | 1.15 | 0.32 | 0.8 - 5.7 | 1 - 1.2 |
|  |  | Female | 3,496 | 1.01 | 1.12 | 0.29 | 0.8 - 3.7 | 1 - 1.1 |
|  |  | Male | 2,701 | 1.1 | 1.19 | 0.36 | 0.8 - 5.7 | 1 - 1.2 |
|  | Liver Transplant | All | 24 | 1.42 | 1.75 | 1.27 | 0.97 - 7.1 | 1.17 - 1.7 |
|  |  | Female | <20 | 1.45 | 2.02 | 1.93 | 0.97 - 7.1 | 1.1 - 1.71 |
|  |  | Male | <20 | 1.4 | 1.58 | 0.67 | 1 - 3.7 | 1.2 - 1.65 |
| Bilirubin | All | All | 35,959 | 0.5 | 0.59 | 0.54 | 0 - 16 | 0.4 - 0.7 |
|  |  | Female | 24,327 | 0.45 | 0.53 | 0.50 | 0 - 16 | 0.35 - 0.6 |
|  |  | Male | 11,632 | 0.6 | 0.71 | 0.60 | 0 - 15.25 | 0.45 - 0.8 |
|  | Control | All | 30,222 | 0.5 | 0.57 | 0.47 | 0 - 16 | 0.4 - 0.7 |
|  |  | Female | 20,803 | 0.45 | 0.52 | 0.45 | 0 - 16 | 0.35 - 0.6 |
|  |  | Male | 9,419 | 0.6 | 0.68 | 0.49 | 0 - 12.6 | 0.45 - 0.8 |
|  | Liver Disease | All | 5,680 | 0.5 | 0.66 | 0.74 | 0.1 - 15.25 | 0.4 - 0.7 |
|  |  | Female | 3,505 | 0.5 | 0.58 | 0.64 | 0.1 - 14.55 | 0.4 - 0.6 |
|  |  | Male | 2,175 | 0.6 | 0.79 | 0.86 | 0.1 - 15.25 | 0.5 - 0.8 |
|  | Liver Transplant | All | 57 | 2.2 | 2.89 | 2.25 | 0.35 - 10.7 | 1.4 - 3.35 |
| **Lab** | **Liver Status** | **Sex** | **N** | **Median** | **Mean** | **SD** | **Range** | **IQR** |
|  |  | Female | <20 | 3.3 | 3.23 | 2.30 | 0.6 - 7.4 | 1.32 - 4.68 |
|  |  | Male | 38 | 2.1 | 2.72 | 2.24 | 0.35 - 10.7 | 1.52 - 2.91 |
| Sodium | All | All | 39,976 | 139 | 138.78 | 2.27 | 122 - 151 | 137.5 - 140 |
|  |  | Female | 26,663 | 139 | 138.81 | 2.22 | 122.5 - 151 | 137.5 - 140 |
|  |  | Male | 13,313 | 139 | 138.72 | 2.37 | 122 - 150 | 137 - 140 |
|  | Control | All | 33,612 | 139 | 138.83 | 2.27 | 122 - 151 | 137.5 - 140 |
|  |  | Female | 22,820 | 139 | 138.83 | 2.23 | 122.5 - 151 | 137.5 - 140 |
|  |  | Male | 10,792 | 139 | 138.83 | 2.35 | 122 - 150 | 137.5 - 140 |
|  | Liver Disease | All | 6,317 | 139 | 138.53 | 2.25 | 123 - 147.5 | 137 - 140 |
|  |  | Female | 3,829 | 139 | 138.72 | 2.15 | 123 - 146 | 137.5 - 140 |
|  |  | Male | 2,488 | 138 | 138.25 | 2.35 | 124 - 147.5 | 137 - 140 |
|  | Liver Transplant | All | 47 | 137 | 136.64 | 3.70 | 127 - 145 | 135 - 138.75 |
|  |  | Female | <20 | 136.5 | 137.04 | 3.46 | 131 - 145 | 135 - 138.88 |
|  |  | Male | 33 | 137 | 136.47 | 3.83 | 127 - 144 | 134.5 - 138.5 |
| MELDNa | All | All | 6,535 | 8.41 | 10.06 | 4.48 | 6.43 - 37.77 | 6.96 - 11.38 |
|  |  | Female | 4,153 | 7.54 | 9.36 | 3.99 | 6.43 - 32.25 | 6.43 - 10.22 |
|  |  | Male | 2,382 | 9.38 | 11.26 | 5.00 | 6.43 - 37.77 | 7.5 - 13.43 |
|  | Control | All | 4,439 | 8.09 | 9.69 | 4.16 | 6.43 - 33.73 | 6.79 - 10.76 |
|  |  | Female | 2,924 | 7.5 | 9.17 | 3.81 | 6.43 - 31.62 | 6.43 - 9.85 |
|  |  | Male | 1,515 | 9.07 | 10.71 | 4.60 | 6.43 - 33.73 | 7.5 - 12.17 |
|  | Liver Disease | All | 2,083 | 8.66 | 10.77 | 4.96 | 6.43 - 37.77 | 7.39 - 12.89 |
|  |  | Female | 1,225 | 8.09 | 9.82 | 4.37 | 6.43 - 32.25 | 6.54 - 10.97 |
|  |  | Male | 858 | 10.31 | 12.14 | 5.42 | 6.43 - 37.77 | 7.76 - 15.1 |
|  | Liver Transplant | All | <20 | 17.31 | 18.35 | 7.08 | 10.12 - 31.01 | 12.01 - 22.37 |
|  |  | Female | <20 | 12.92 | 13.76 | 2.65 | 11.72 - 17.47 | 11.94 - 14.74 |
|  |  | Male | <20 | 20.79 | 20.39 | 7.57 | 10.12 - 31.01 | 16.76 - 25.39 |

Supplementary Table 7. Sex differences in median MELDNa component labs and calculated MELDNa stratified by liver status. Differences were assessed using a Student’s t-test.

| **Lab** | **Group** | **VUMC p-value** | **All of Us p-value** |
| --- | --- | --- | --- |
| Creatinine | All | <2.22 x 10^-308^ | <2.22 x 10^-308^ |
|  | Control | <2.22 x 10^-308^ | 1.22 x 10^-316^ |
|  | Liver Disease | 1.03 x 10^-291^ | 8.39 x 10^-34^ |
|  | Liver Transplant | 0.001 | 0.9606 |
| INR | All | 1.47 x 10^-128^ | 8.48 x 10^-40^ |
|  | Control | 2.65 x 10^-114^ | 3.49 x 10^-31^ |
|  | Liver Disease | 1.80 x 10^-14^ | 1.43 x 10^-09^ |
|  | Liver Transplant | 0.738 | 0.528 |
| Bilirubin | All | <2.22 x 10^-308^ | 5.11 x 10^-169^ |
|  | Control | <2.22 x 10^-308^ | 4.44 x 10^-168^ |
|  | Liver Disease | 6.42 x 10^-54^ | 8.64 x 10^-13^ |
|  | Liver Transplant | 0.262 | 0.317 |
| Sodium | All | <2.22 x 10^-308^ | 4.08 x 10^-03^ |
|  | Control | 0.599 | 0.6425 |
|  | Liver Disease | 1.88 x 10^-23^ | 3.16 x 10^-09^ |
|  | Liver Transplant | 0.364 | 0.565 |
| MELDNa | All | <2.22 x 10^-308^ | 3.31 x 10^-48^ |
|  | Control | 3.16 x 10^-313^ | 4.64 x 10^-33^ |
|  | Liver Disease | 2.20 x 10^-54^ | 1.87 x 10^-14^ |

Supplementary Table 8. Sex differences in maximum MELDNa component labs and calculated MELDNa stratified by liver status in VUMC. Statistical significance was assessed using a Student’s t-test.

| Lab | Group | p-value |
| --- | --- | --- |
| Creatinine | All | <2.22 x 10^-308^ |
|  | Control | <2.22 x 10^-308^ |
|  | Liver Disease | 5.19 x 10^-125^ |
|  | Liver Transplant | 0.047 |
| INR | All | 7.11 x 10^-55^ |
|  | Control | 4.17 x 10^-49^ |
|  | Liver Disease | 1.00 x 10^-05^ |
|  | Liver Transplant | 0.198 |
| Bilirubin | All | <2.22 x 10^-308^ |
|  | Control | <2.22 x 10^-308^ |
|  | Liver Disease | 3.71 x 10^-40^ |
|  | Liver Transplant | 0.026 |
| Sodium | All | 1.19 x 10^-195^ |
|  | Control | 2.41 x 10^-177^ |
|  | Liver Disease | 0.041 |
|  | Liver Transplant | 0.021 |
| MELDNa | All | 8.51 x 10^-304^ |
|  | Control | 6.41 x 10^-258^ |
|  | Liver Disease | 2.33 x 10^-46^ |
|  | Liver Transplant | 0.186 |

Supplementary Table 9. Sex differences in average decompensation counts stratified by sex and liver status. Statistical significance was assessed using Student’s t-tests.

| **Liver Status** | **Sample** | **Female Average (SD)** | **Male Average (SD)** | **p-value** |
| --- | --- | --- | --- | --- |
| All | VUMC | 0.019 (0.16) | 0.027 (0.20) | 2.20 x 10^-73^ |
|  | All of Us | 0.008 (0.10) | 0.012 0.13() | 1.31 x 10^-16^ |
| Controls | VUMC | 0.011 (0.11) | 0.144 (0.13) | 2.86 x 10^-31^ |
|  | All of Us | 0.004 (0.07) | 0.006 (0.08) | 5.76 x 10^-06^ |
| Liver Disease | VUMC | 0.206 (0.56) | 0.253 (0.61) | 2.75 x 10^-10^ |
|  | All of Us | 0.094 (0.36) | 0.135 (0.46) | 3.15 x 10^-06^ |
| Liver Transplant | VUMC | 1.599 (1.09) | 1.341 (1.11) | 0.00481 |
|  | All of Us | 1.100 (0.74) | 1.222 (0.94) | 0.708 |
